# AI-Enhanced Reconstruction of the 12-Lead Electrocardiogram via 3-Leads with Accurate Clinical Assessment

**DOI:** 10.1101/2024.01.30.24302001

**Authors:** Federico Mason, Amitabh C. Pandey, Matteo Gadaleta, Eric J. Topol, Evan D. Muse, Giorgio Quer

**Author notes:** Correspondence to: Giorgio Quer, Scripps Research Translational Institute, 3344 N Torrey Pines Ct Plaza Level, La Jolla, 92037, California, USA;, Evan D. Muse, Scripps Research Translational Institute, 3344 N Torrey Pines, Suite 300, La Jolla, 92037, California, USA. These authors contributed equally: F.M. and A.C.P.

## Abstract

The 12-lead electrocardiogram (ECG) is an integral component to the diagnosis of a multitude of cardiovascular conditions. It is performed using a complex set of skin surface electrodes, limiting its use outside traditional clinical settings. We developed an artificial intelligence algorithm, trained over 600,000 clinically acquired ECGs, to explore whether fewer leads as input are sufficient to reconstruct a full 12-lead ECG. Two limb leads (I and II) and one precordial lead (V3) were required to generate a reconstructed synthetic 12-lead ECG highly correlated with the original ECG. An automatic algorithm for detection of acute myocardial infarction (MI) performed similarly for original and reconstructed ECGs (AUC=0.94). When interpreted by cardiologists, reconstructed ECGs achieved an accuracy of 81.4±5.0% in identifying ST elevation MI, comparable with the original 12-lead ECGs (accuracy 84.6±4.6%). These results will impact development efforts to innovate ECG acquisition methods with simplified tools in non-specialized settings.

## INTRODUCTION

With over 300 million being performed worldwide on an annual basis,^1^ the 12-lead electrocardiogram (ECG) has established itself as a bedrock diagnostic in the assessment of cardiovascular disease.^2-5^ Using a complicated array of 10 individual skin-surface electrodes, a series of 12 individual electrical vectors is arranged to assist in the diagnosis of an array of cardiopulmonary diseases. However, the acquisition of a 12-lead ECG has not iterated to great degree from its initial inception. It can be cumbersome, requiring special equipment available only at a hospital or clinic, and specially trained individuals to perform and interpret the ECG. Over the last several years, technological advancements made it possible to monitor specific cardiac activity through wearable devices including smart watches, patch monitors and apps with improved quality and speed. However, ECG monitoring in this setting is often limited to a single lead (typically lead I) or few limb leads, which are inadequate for confidently diagnosing abnormalities limited to specific myocardial regions, such as acute myocardial infarction (MI).^6^ Since the specific patterns suggestive of an acute MI may be reflected in the limb leads, the precordial leads, or a combination of limb and precordial leads, current guidelines require the use of a 12-lead standard ECG for clinical interpretation.

The 12 leads in a standard ECG are not fully independent and are known to be in-part correlated,^7^ thus over the last 30 years techniques have been proposed to synthesize a full standardized ECG from a limited lead set.^8^ While initial advancements in this field relied on linear transformation models, the diffusion of artificial intelligence (AI) enabled the development of more sophisticated approaches.

Prior studies have primarily relied on patient-specific models^9^ or have been derived from limited datasets,^10,11^ potentially limiting their generalizability. In this study, our aim was the development of a reconstruction algorithm with the purpose of synthesizing a complete 12-lead ECG from a limited subset of leads. To this end, we leveraged a large retrospective dataset of clinically obtained 12-lead ECGs. Additionally, we assessed the clinical utility of this reconstructed ECG involving three cardiologists, using ST-elevation MI (STEMI) as a case study.

## RESULTS

### Dataset curation and categorization

We considered a working dataset with 618,880 ECGs from 274,738 unique individuals. In particular, 46.19% of the ECGs were recorded from individuals 18-60 years old, 20.22% are associated with non-Caucasian individuals, and 50.44% were from female individuals. Normal sinus rhythm was present in 57.01% of the ECGs, while the rest was characterized by some form of arrythmia, with sinus, atrial, and ventricular arrhythmias present in 26.08%, 17.08%, and 7.24% of the ECGs, respectively. Cardiac conduction disorders were present in 30.66% of the ECGs, while 25.88% presented a repolarization abnormality in the ST segment or T wave. Ventricular hypertrophy, deviations of the cardiac axis and ischemia were present in 7.55%, 14.71%, and 10.26% of the ECGs, respectively. (Table 1)

**Table 1.** Reconstruction and classification performance according to the demographic and clinical features of the dataset. The table describes the system performance associated with different classes of data, depending on demographic characteristics of the population and features of the ECGs. For each class, the table reports total number of individuals, total number of ECGs, and percentage of ECGs associated with acute MI. The reconstruction performance is assessed in terms of coefficient of determination (R2), and mean squared error (MSE), while the classification performance (MI identification) is assessed in terms of area under the curve (AUC) or the receiver operating characteristic (ROC). The confidence intervals for R2 and MSE were computed considering a confidence level of 95%.

| | | Number of elements in the overall dataset | | | R2 [%] in the test set (average ± confidence interval) | | MSE [mV<sup>2</sup>] in the test set (average ± confidence interval) | | Area Under the Curve (AUC) in the test set | | |
| --- | --- | --- | --- | --- | --- | --- | --- | --- | --- | --- | --- |
|  |  | Number of individuals | Number of ECGs | Percentage of acute MI | Using I+II | Using I+II+V3 | Using I+II | Using I+II+V3 | Using I+II | Using I+II+V3 | Using 12-leads |
| | Total | 274738 | 618880 | 2.94 | 52.54 ± 0.19 | 72.62 ± 0.13 | 0.0253 ± 0.0002 | 0.0126 ± 0.0001 | 0.92 | 0.94 | 0.94 |
| Demographic features | 18-60 years old | 126902 | 208795 | 3.71 | 52.57 ± 0.34 | 72.62 ± 0.23 | 0.0252 ± 0.004 | 0.0126 ± 0.0002 | 0.91 | 0.94 | 0.95 |
| | 60-80 years old | 120787 | 269483 | 2.79 | 52.38 ± 0.28 | 72.59 ± 0.20 | 0.0254 ± 0.004 | 0.0125 ± 0.0002 | 0.92 | 0.94 | 0.94 |
| | More than 80 years old | 53092 | 140602 | 2.09 | 52.79 ± 0.38 | 72.67 ± 0.28 | 0.0254 ± 0.0005 | 0.0127 ± 0.0003 | 0.91 | 0.95 | 0.95 |
| | Female | 138567 | 295049 | 1.81 | 52.43 ± 0.27 | 72.59 ± 0.19 | 0.0255 ± 0.0003 | 0.0127 ± 0.0002 | 0.91 | 0.94 | 0.94 |
| | Male | 136370 | 323817 | 3.98 | 52.63 ± 0.25 | 72.65 ± 0.19 | 0.0252 ± 0.0003 | 0.0125 ± 0.0002 | 0.92 | 0.94 | 0.94 |
| | Caucasian | 207472 | 477763 | 2.68 | 52.56 ± 0.21 | 72.62 ± 0.15 | 0.0253 ± 0.0003 | 0.0126 ± 0.0001 | 0.92 | 0.94 | 0.94 |
| | Non-Caucasian | 55564 | 103664 | 4.20 | 52.24 ± 0.46 | 72.72 ± 0.32 | 0.0250 ± 0.0006 | 0.0128 ± 0.0003 | 0.91 | 0.95 | 0.94 |
| Rhythm features | Normal sinus rhythm | 218589 | 352849 | 2.86 | 52.59 ± 0.24 | 72.62 ± 0.17 | 0.0251 ± 0.0003 | 0.0126 ± 0.0002 | 0.92 | 0.94 | 0.94 |
| | Sinus arrhythmia | 142354 | 161373 | 3.27 | 52.37 ± 0.38 | 72.51 ± 0.27 | 0.0257 ± 0.0005 | 0.0125 ± 0.0002 | 0.92 | 0.94 | 0.95 |
| | Atrial arrhythmia | 69295 | 105720 | 2.53 | 52.59 ± 0.44 | 72.58 ± 0.34 | 0.0253 ± 0.0006 | 0.0127 ± 0.0003 | 0.91 | 0.93 | 0.93 |
| | Ventricular arrhythmia | 44664 | 44830 | 2.96 | 52.80 ± 0.69 | 72.68 ± 0.58 | 0.0252 ± 0.0008 | 0.0126 ± 0.0005 | 0.94 | 0.93 | 0.93 |
| Morphological features | Conduction disorders | 113936 | 189766 | 3.03 | 52.51 ± 0.33 | 72.57 ± 0.24 | 0.0254 ± 0.0004 | 0.0126 ± 0.0002 | 0.91 | 0.94 | 0.94 |
| | Repolarization abnormality | 114233 | 160170 | 7.62 | 52.50 ± 0.36 | 72.45 ± 0.28 | 0.0253 ± 0.0004 | 0.0126 ± 0.0002 | 0.91 | 0.95 | 0.94 |
| | Cardiac hypertrophy | 44387 | 46730 | 3.78 | 52.49 ± 0.66 | 72.40 ± 0.51 | 0.0248 ± 0.0008 | 0.0127 ± 0.0004 | 0.92 | 0.95 | 0.95 |
| | Axis deviation | 56787 | 91060 | 2.85 | 52.72 ± 0.54 | 72.51 ± 0.32 | 0.0245 ± 0.0006 | 0.0127 ± 0.0003 | 0.90 | 0.94 | 0.95 |
| | Ischemia | 48877 | 63486 | 3.59 | 52.42 ± 0.57 | 72.54 ± 0.48 | 0.0255 ± 0.0007 | 0.0127 ± 0.0004 | 0.92 | 0.94 | 0.95 |
| | Prior infarct | 117135 | 293277 | 2.11 | 52.28 ± 0.29 | 72.61 ± 0.20 | 0.0256 ± 0.0003 | 0.0126 ± 0.0002 | 0.92 | 0.94 | 0.94 |

Focusing on the subset of ECGs associated with myocardial infarction (MI), 47.40% of the data had signs of past MI, while 18,215 ECGs (2.94%) present evidence of acute MI. Regarding the anatomical location of the acute MI, we had 8.25% anterior, 0.37% septal, 5.72% lateral, 5.33% anteroseptal, 6.29% anterolateral, 4.08% inferolateral, 32.15% inferior/posterior, and 40.44% unspecified.

### 12-lead reconstruction algorithm

The reconstruction algorithm’s performance was assessed in terms of mean squared error (MSE) and coefficient of determination (R2) between the original precordial leads and those synthesized by the algorithm. When the algorithm used only two limb leads as input, performance was relatively poor with MSE = 0.0253 ± 0.0002 mV^2^ and R2 = 52.54 ± 0.19 %. Adding a single precordial lead in input significantly improved the reconstruction accuracy, with the best performance observed using lead V3, MSE = 0.0126 ± 0.0001 mV^2^ and R2 = 72.62 ± 0.13%. Replacing lead V3 with either lead V2 or lead V4 resulted in a slight decrease in reconstruction accuracy. (Figure 1)

**Figure 1.**
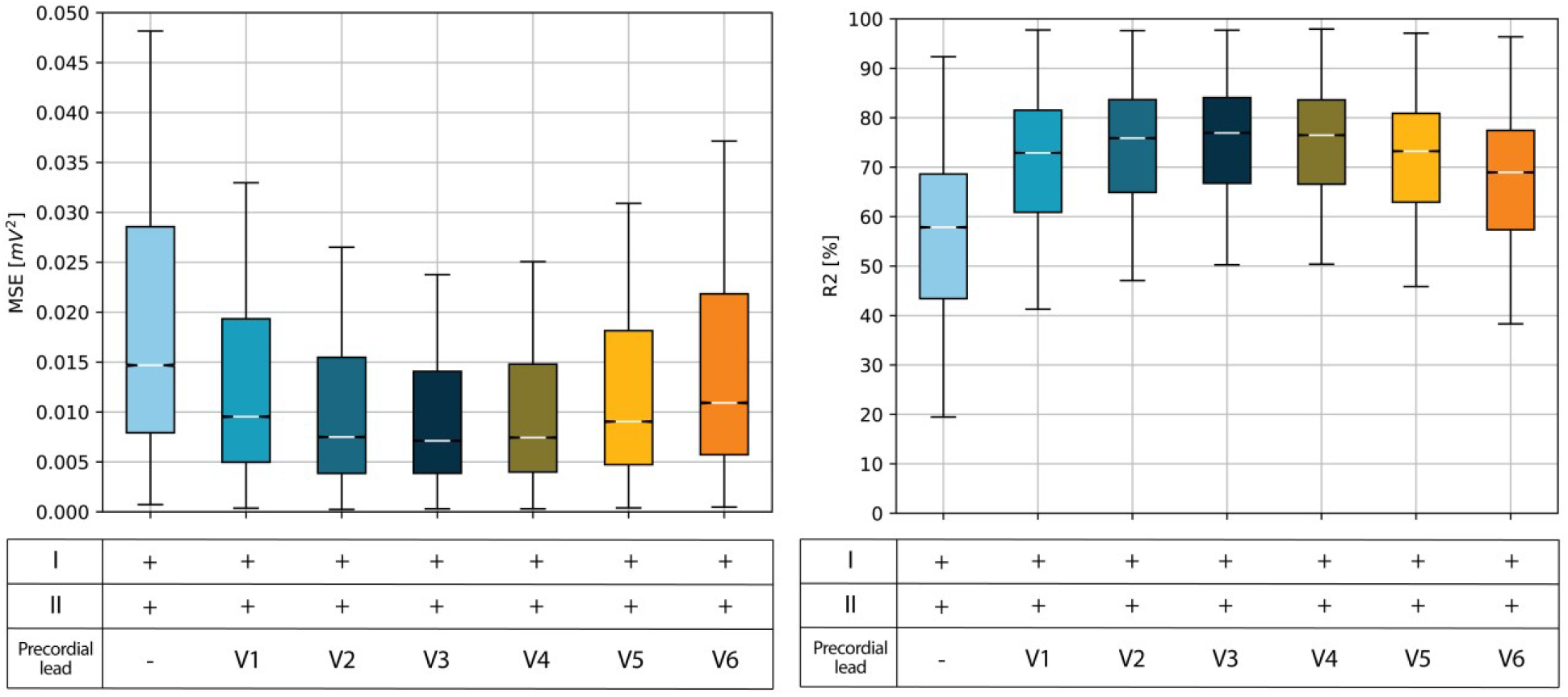
Distributions of mean square error and coefficient of determination in 12-lead ECG reconstruction. Boxplot of the mean squared error (MSE) and the coefficient of determination (R2) distribution according to various input configurations for the reconstruction of a 12-lead ECG. The white line in the middle of each box represents the distribution median, the box edges are the 25th and 75th percentiles, while the box whiskers are the 5th and 95th percentiles. The MSE is given by the sum of the squared difference between the original ECG values and those reconstructed by the designed algorithm. Instead, the R2 represents the fraction of variance of the original ECGs captured by the reconstruction model and is independent of the actual scale of the data.

### Classification using the reconstructed signal

The classification algorithm’s accuracy was evaluated using three distinct versions of 12-lead ECG, obtained using as input: the original 12-lead ECG (Original), the 12-lead ECG reconstructed from two limb leads (I+II), and the 12-lead ECG reconstructed from limb leads and precordial lead (I+II+V3). The area under the operating characteristic curve (AUC) for the classification algorithm with input I+II+V3 was AUC=0.94, which is equivalent to the performance obtained using the original 12-lead ECG, while the AUC with the I+II version as input was considerably lower. (Figure 2)

**Figure 2.**
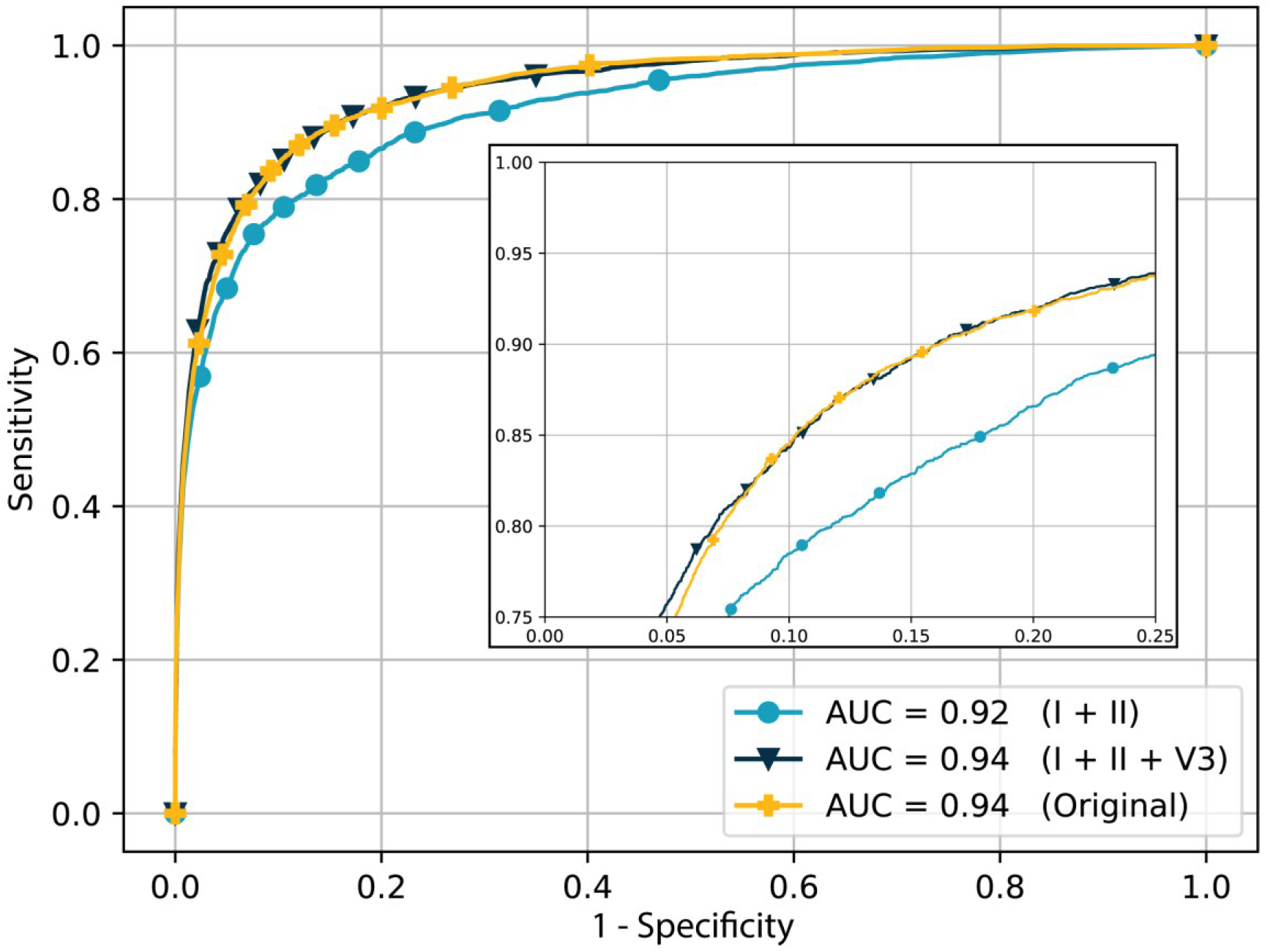
Receiver operating characteristics curves for acute MI detection. Receiver operating characteristic (ROC) curves for acute MI detection according to various input configurations. The ROC curve depicts the performance of the detection system while varying the discrimination threshold between sensitivity and specificity. In our case, the sensitivity, also known as true positive ratio, corresponds to the probability that an ECG is diagnosed as acute MI, conditioned on the fact the original signal was labelled as acute MI. Instead, the specificity, also known as true negative ratio, corresponds to the probability that an ECG is not diagnosed as acute MI, conditioned on the fact that original signal was not labelled as acute MI.

Performance of both reconstruction and classification algorithms are reported for all the demographic and clinical features described. (Table 1)

### Clinical assessment of the reconstruction algorithm

The cardiologists involved in the clinical interpretation of our framework were able to correctly discriminate between the presence or absence of ST elevation MI (STEMI) in 84.6±4.6% of the cases when using the original ECGs, 81.4±5.0% of cases when using the I+II+V3 version, and 75.5±5.5% of cases when using the I+II version, showcasing the importance of one precordial lead for the reconstruction. The specificity remained consistently at 100% in all cases. This means that all ECGs identified as STEMI, including those generated synthetically, were confirmed as STEMI cases. The sensitivity was 68.7±8.5% for the original 12-lead ECG, 62.4±8.8% for the synthetized 12-lead ECG with I+II+V3, and 51.3±9.0% for the synthetized 12-lead ECG with I+II. (Figure 3) The results showed that the ability to identify STEMI from a synthesized 12-lead ECG (I+II+V3) is not inferior to the one obtained from the original ECG with a margin of error of 10% (p-value=0.026).

**Figure 3.**
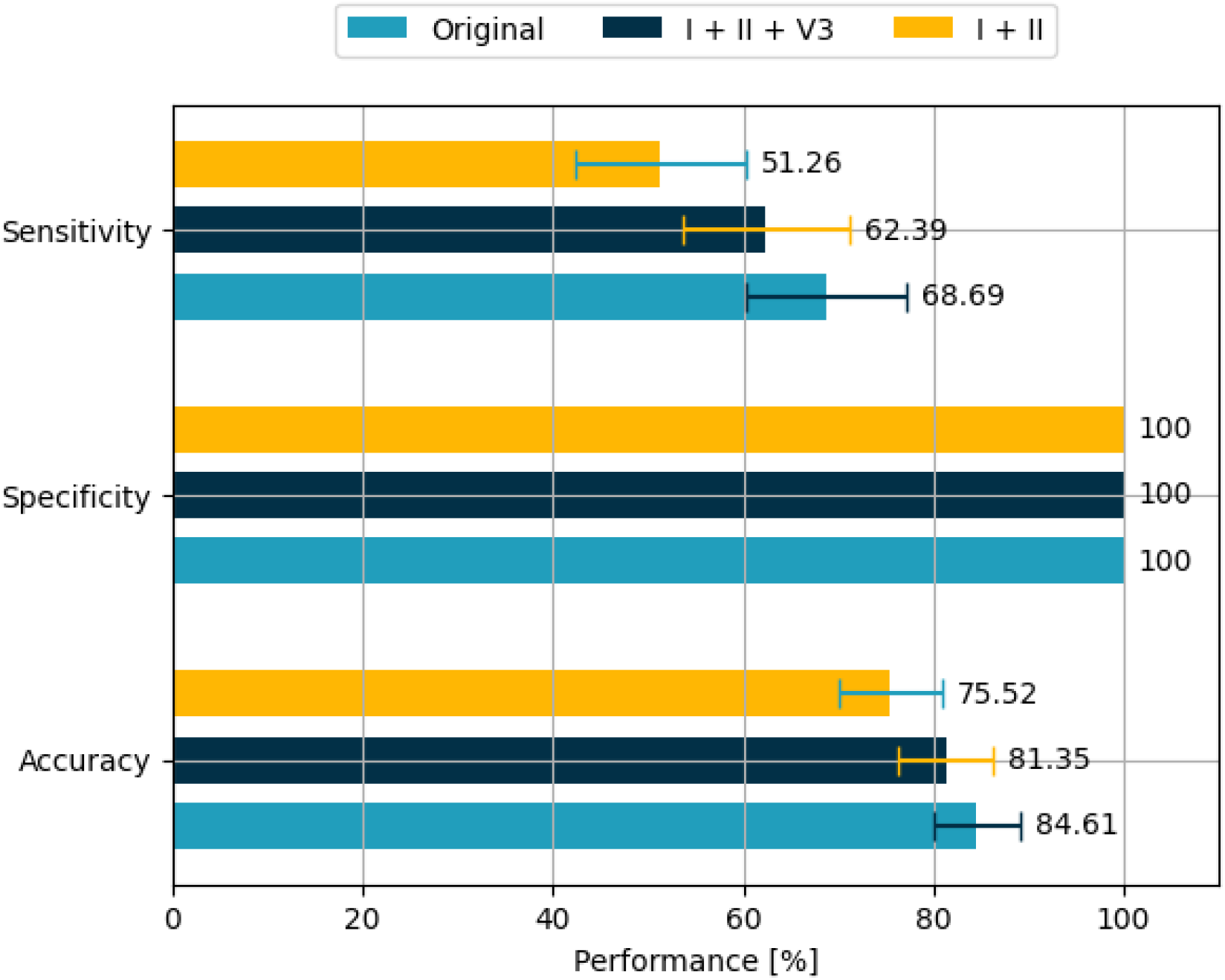
Detection accuracy of the original and reconstructed ECGs in clinical validation. Sensitivity, specificity and accuracy obtained during the clinical validation of the proposed reconstruction model. The sensitivity, also known as true positive ratio, corresponds to the probability that an ECG is diagnosed as STEMI, conditioned on the fact the original signal presents STEMI evidence. The specificity, also known as true negative ratio, corresponds to the probability that an ECG is diagnosed as not-STEMI, conditioned on the fact that the original signal does not present STEMI evidence. Finally, the accuracy is given by the ratio between the number of correct diagnoses and the total number of ECGs analyzed

## DISCUSSION

In the present study, we designed a novel AI algorithm for the reconstruction of a full 12-lead ECG from two limb leads only (I and II) or two limb leads and a single precordial lead (I, II, and V3). We assessed the accuracy of the reconstructed 12-lead ECG by using an automatic detection model, trained to classify ECG records according to the evidence of acute MI. The detection model was equally effective with input of an original 12-lead or reconstructed (I+II+V3) ECG. This result provides initial evidence that I+II+V3 leads may be sufficient for reconstructing a 12-lead ECG towards the identification of acute MI.

We also showed that the ECGs reconstructed by our AI algorithm can be effectively interpreted by a cardiologist for diagnosing STEMI, with a limited performance reduction with respect to the standard 12-lead ECGs. In this test, as seen in earlier findings,^14^ sensitivity was relatively low for both the original and reconstructed 12-lead ECGs. This could be attributed to the cardiologists’ assessment of the ECGs without additional information about the patient and their lack of prior knowledge regarding the elevated prevalence of STEMI cases in the dataset (50%). Consequently, they might have been more cautious in diagnosing these ECGs as STEMI. While larger multi-site clinical trials are needed to confirm this initial evidence, these results are a promising step towards the use of this algorithm when a 12-lead ECG is not available.

This study builds on our previous work that proposed an AI architecture for the analysis of single-lead ECGs.^15,16^ Several approaches in the literature attempted to reconstruct a full 12-lead ECG, e.g., leveraging the correlation among different leads included using linear transformation matrices,^8,17^ or temporal-based models.^18,19^ However, most of these previous works designed an individualized algorithm,^20^ which, in turn, limits the effectiveness of the proposed application. Lead interdependency varies from individual to individual and, thus, more advanced models are needed to shape the relation between limb and precordial leads. Herein lies the strength of supervised techniques that can approximate complex functions by learning from a large amount of labeled data, enabling the definition of new tools for synthetizing ECGs from partial information. A first example of AI for ECG reconstruction exploited a feed-forward neural network (FNN) system to generate a full 12-lead ECG using the limb leads combined with V2 as input.^9^ More sophisticated techniques were also proposed, including convolutional neural networks (CNNs)^10^ and long short term memory (LSTM) models,^11^ both suitable tools for processing time-series like an ECG signal. Notably, CNNs provided high-quality results for the identification of atrial fibrillation and other rhythm-related abnormalities,^21^ or for the automatic detection of STEMI from limb leads only.^22^

Despite these solutions to identify heart diseases in an automatic manner,^23-25^ the reconstruction of a 12-lead ECG represents a fundamental step towards detection of acute coronary syndrome, specifically STEMI, that can be effectively verified by a cardiologist. Previous research that exploited AI for this goal was based on limited-size datasets (a few hundred records), not suitable for the training of complex learning models.^9,10,26^ In other studies, data from the same patient was used both in training and testing, affecting generalizability.^11,27^

Our results suggest that it is possible to reconstruct a 12-lead ECG from the measurement of a limited set of 1 precordial and 2 limb leads, and that the synthetized signal can be used by a cardiologist for the detection of STEMI. While most of previous investigations assumed that septal lead V2 is the most important precordial lead,^8^ we have shown that measuring anterior lead V3 provides the best accuracy for the reconstruction. This finding may be attributed to the central position of V3, resulting in stronger correlations not only with anterior lead V4 but also with septal and lateral leads. Our study highlighted how the reconstructed 12-lead ECG is not only useful for an automatic algorithm in the identification of acute MI, but it can also be interpreted by cardiologists and used to identify STEMI. Considering that the required input can be obtained using commercial sensors without the need for a complete 12-lead ECG, this solution becomes particularly valuable in scenarios where acquiring a full 12-lead ECG is impractical. This is especially relevant in settings such as care facilities with limited resources or remote locations lacking clinical infrastructures. In these scenarios, the proposed system could facilitate early diagnosis of acute MI, potentially reducing the time for medical intervention.

This work (and similar studies) represents a step towards the transitioning from brick-and-mortar clinical facilities to remote, direct-to-participant healthcare accessible to everyone. This is now possible using more accurate sensor technologies, the use of AI algorithms to learn from massive datasets and reproduce clinical-level signals, and the ubiquitous connectivity that enables rapid and constant two-way communication from remote locations to the clinic.^28-30^ The potential of digital technologies lies in their capacity to bring healthcare closer to individuals at any time, even when accessing a clinical facility may be challenging.

Using a limited number of leads to capture the essential information of a 12-lead ECG has the potential to facilitate the diagnosis of ischemia, arrhythmias, and other heart-related conditions. Solutions like the one proposed in this study may enable medical examinations, such as cardiac stress tests, to be performed in a home setting, making the health system more agile, especially in combination with the other possibilities offered by telemedicine. The reconstruction of a 12-lead ECG may serve as a valuable tool also in a hospital setting, preempting the need for a technician, reducing the time required to record the standard ECG leads and offering a preliminary diagnosis procedure during emergency room admissions or ambulance transports.

### Limitations

The designed algorithm is based on data recorded through conventional 12-lead ECGs, where only lead I, II and V3 are considered as input of the reconstruction algorithm. The measurements are indeed performed by highly trained clinical personnel in a hospital system, thus the challenges linked to different recording systems, potentially more prone to inaccuracies, performed by non-clinical personal outside of a clinical setting, are yet to be evaluated. Furthermore, the dataset is sourced exclusively from a single hospital system, with a good gender balance (percentage of female is 50.44%) but a higher representation of Caucasian individuals (with a ratio of 4:1 compared to non-Caucasian individuals). The rate of ECGs diagnosed with acute MI is higher among non-Caucasian individuals (4.20%) in comparison to Caucasian individuals (2.68%). Although there is currently no substantial evidence indicating significant disparities in ECG interpretation based on race, further investigation with data from individuals with diverse demographic characteristics is needed to exclude this potential bias. Finally, the algorithm should be evaluated in a multi-site study, where different ECG systems are included, and potentially it should be re-trained accordingly.

### Conclusions

These results illustrate the fidelity of a fully reconstructed 12-lead ECG using two limb leads (I and II) and one precordial lead (V3), which could be collected using a simple mobile sensing platform, promoting future innovation. Such algorithm and technology may be used outside of a clinical setting, allowing for time-sensitive STEMI diagnoses, thereby potentially facilitating prompt emergency procedure.

## METHODS

### Study population

This work was based on an initial dataset of 1,718,909 de-identified ECG performed in-clinic, collected from 554,120 unique individuals from 2008 to 2019 using the Scripps Health GE MUSE system (GE Healthcare, Chicago, IL). All ECGs were exported in their raw form and processed in a standardized manner for the retrospective analysis. The ECGs from individuals younger than 18 years or associated with recording errors were excluded. For the training and evaluation of all the algorithms, we considered a subset of the data, namely the working dataset, with 618,880 ECGs, which were obtained considering all available ECGs with a diagnosis of MI in the ECG (n=309,440), and an equal number of non-MI ECGs selected at random.

### Ethical considerations

The protocol for this project was reviewed and approved by the Scripps Office for the Protection of Research Subjects (IRB-20-7504).

### Dataset description

Each element in the dataset was composed of 12 electrical signals of length 10 seconds, recorded with a sampling frequency of 250 or 500 Hz. All signals were resampled at a frequency of 250 Hz. The ECG leads were divided into two classes: limb (I, II, III, aVR, aVL, aVF) and precordial (V1, V2, V3, V4, V5, V6) leads. Limb leads are linearly dependent, all six limb leads can be obtained by measuring only two of them.^29^ The relationships between limb leads are detailed in the following equations.

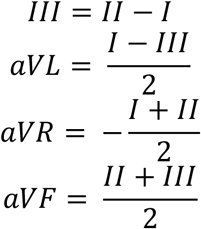

Each element of the dataset was associated with an automatic diagnosis, revised by the clinician that analyzed and finalized the ECG recording. These final diagnoses were utilized to run a text mining algorithm and define the clinical features of each element. These features described the characteristics of the ECG signal, considering both the cardiac conduction and features consistent with structural and clinical diagnoses.

We divided the working dataset into three mutually exclusive subsets, which were then used as training (n=433,554), validation (n=46,371), and testing (n=46,292) data for our analysis. The proportion between MI and non-MI labels was maintained in each subset, ensuring that approximately half of the ECGs used for the training were associated with evidence of MI. Moreover, in cases where more than one ECG was associated with the same individual, all of the ECGs for that individual were grouped into the same subset.

### Algorithm architecture

The main goal of this work was the design of a reconstruction algorithm to produce a 12-lead ECG from a subset of the signal leads. The algorithm was based on an AI architecture leveraging residual convolutional neural networks (RCNNs). The architecture could take any combination of the signal leads, whether limb or precordial, as input, while consistently producing the full 12-lead ECG as output. The input leads were processed according to a feature scaling approach,^30^ which preserves the mathematical differences among different data. Each lead was mapped in the interval between 0 and 1, in order to facilitate the learning phase of the algorithm. We considered a minimum and maximum value of -2.5 and 2.5 mV for each lead, where values larger than 2.5 mV were mapped to 1, while values lower than -2.5 mV were mapped to 0.

The algorithm architecture was organized into three sections. The first included three independent RCNN blocks, each taking a single lead as input and returning the lead features as output, which were encoded in multi-dimensional vectors of 2500 samples and 32 channels. In the second section, the lead features were aggregated and processed through a RCNN block returning a multi-dimensional vector of 2500 samples and 192 channels as output. Finally, the third section included six independent blocks, each receiving a copy of the aggregated features and computing one of the six precordial leads of the reconstructed ECG. (Figure 4)

**Figure 4.**
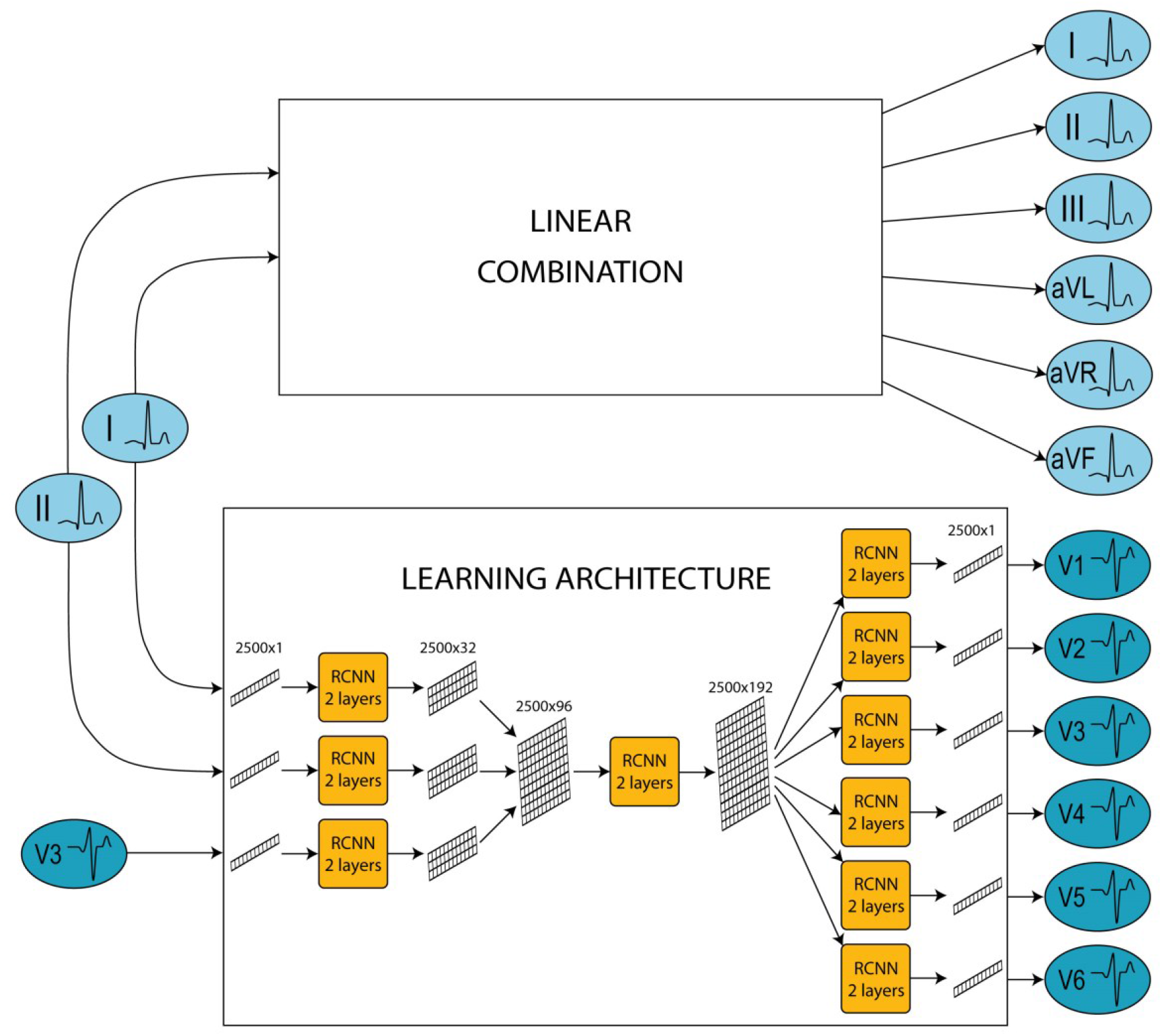
12-lead ECG reconstruction algorithm architecture. The architecture of the 12-lead ECG reconstruction algorithm includes two modules. The upper module takes limb I and II as input and exploits a linear combination to generate the limb leads. The lower module takes lead I, II and V3 as input, and exploits a deep Neural Network (NN) architecture to generate the precordial leads. Particularly, the NN architecture is organized into three different sections, each including a different number of NN blocks. The first section includes three RCNN blocks with the function of extracting the features of each of the input leads. The second section includes a single RCNN block receiving the aggregated lead feature as input and returning a unique multi-dimensional vector as output. In the last section, the same vector is processed through six independent RCNN blocks, each of which returns a different precordial lead of the reconstructed signal. The aggregated output of the two modules returns the 12-lead ECG.

To evaluate the accuracy of the reconstructed signals, we designed a separate classification algorithm to automatically detect acute MI from a 12-lead ECG. The architecture of the classification algorithm received eight leads (I+II and V1-V6) as input. Each lead was encoded as a vector of 2500 samples, with values normalized between 0 and 1. In the first step of the architecture, these vectors were processed individually, obtaining eight multi-dimensional vectors of 2500 samples and 32 channels. In the second section, features were aggregated and processed through a single RCNN block returning a single-dimensional vector of 128 values. In the last section, a feedforward neural network transformed this vector into a single scalar value, which is processed through a sigmoid function. The final output represented the probability of acute MI. (Figure 5)

**Figure 5.**
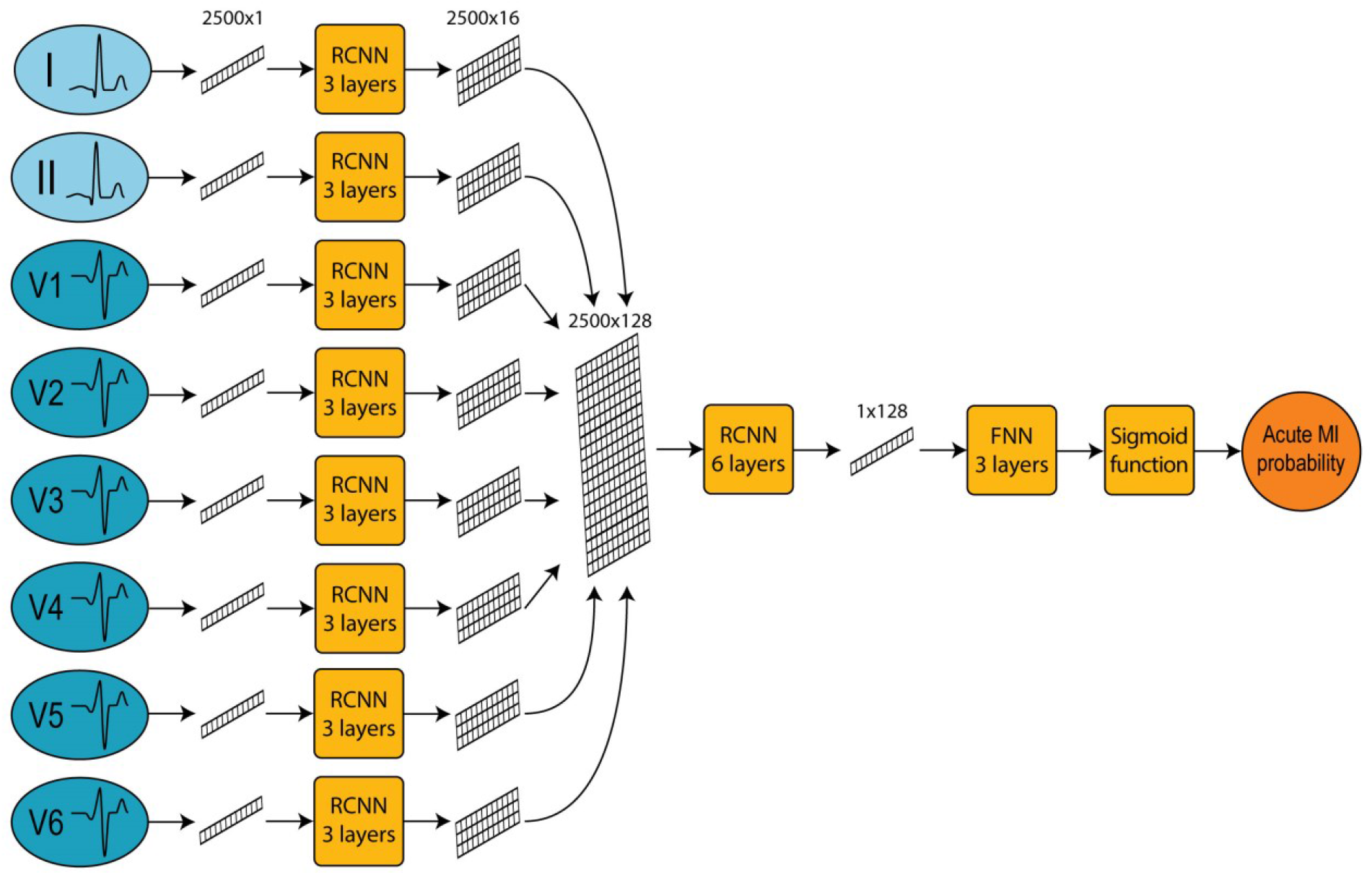
Acute MI detection algorithm architecture. The architecture of the acute MI detection algorithm is organized into three different sections. The first section includes eight recurrent convolutional neural network (RCNN) blocks extracting the features of each of the eight input leads. The second section includes a single RCNN block receiving the aggregated lead feature as input and returning a unique multi-dimensional vector as output. In the last section, the vector obtained is processed through a Feed-Forward Neural Network (FNN) and, finally, a sigmoid function, returning the probability of detecting an acute MI.

### Algorithm training and evaluation

During the training phase, the reconstruction algorithm was encouraged to minimize the mathematical distance between the original precordial leads and the precordial leads generated by the learning architecture, considering R2 as the loss function. During the testing phase, we assessed the performance of the reconstruction algorithm in terms of MSE and R2. The MSE quantified the average squared difference between the original precordial leads and those reconstructed with our approach. Instead, R2 was utilized to assess how effectively the reconstructed signal captures the variance of the original signal, regardless of its amplitude. In the case of a perfect reconstruction, we would obtain MSE=0 and R2=100%.

The goal of the classification algorithm was to associate each ECG to a label denoting the presence or absence of acute MI. Hence, during the training phase, the algorithm was trained to minimize the cross-entropy (CE) between the true labels of the data and the labels predicted by the learning architecture. During the testing phase, we evaluated the performance of the classification algorithm by calculating the AUC for each input configuration.

### Cardiologist interpretation of the reconstructed ECGs

To assess the interpretability of the reconstructed signal, three board-certified cardiologists were asked to analyze a set of multiple ECGs, including both original and synthetized signals, identifying the ECG consistent with a STEMI diagnosis. In determining the sample size for the clinical interpretation of our system, we hypothesized that a cardiologist’s accuracy in assessing the original ECGs and the signals reconstructed from the limb leads was 95% and 90%, respectively. Therefore, we considered a sample of 238 ECGs, with 119 of them showing indications of STEMI, while the remaining 119 comprised normal ECGs or exhibited other non MI abnormalities. The correct diagnoses of these data were verified by two expert cardiologists (not involved in the test) before the test, who analyzed all the ECGs and agreed on the correct label.

Using our reconstruction algorithm, we generated three versions for each of the selected ECGs. The first version was the original 12-lead ECG (Original), the second version was the 12-lead ECG synthetized by our reconstruction algorithm considering two limb leads (I+II) as input, and the third version was the synthetized 12-lead ECG considering limb leads and precordial lead (I+II+V3) as input. For each ECG, each of the three versions (I+II, I+II+V3, and Original) was randomly assigned to one of the three cardiologists, so that each cardiologist was evaluating 238 ECGs, without knowing which of them were original 12-lead ECG, and which were synthetized. The ECGs were presented one by one using MyDataHelps, an online platform provided by CareEvolution. After enrolling to the platform, each cardiologist was asked to examine the 238 assigned ECGs, associating to each signal a diagnosis among: “STEMI”, “non-STEMI”, and “Unable to determine”. The cardiologist could complete the test at their own pace, interrupting it at their own will.

Comparing the answers of the cardiologists with the original data labels, we estimated the detection accuracy, sensitivity, and specificity, associated with each input configuration of the reconstruction algorithm (I+II, I+II+V3, and Original). We then proved the non-inferiority of the I+II+V3 system with respect to the original ECG using an unpooled z-test and considering 10% as margin of error. We considered the z-test’s outcome statistically significant if associated with a p-value smaller than 0.05. The p-value is the probability of observing a given event given that the null hypothesis is true; in our work, the null hypothesis is that the accuracy of the I+II+V3 system is more than 10% lower than that obtained when using the original ECGs as input. Hence, a smaller the p-value corresponds to a stronger evidence that I+II+V3 does not lead to relevant performance degradation.

### DATA SHARING

All code used to develop the deep learning algorithm can be requested by contacting the corresponding author. This study is retrospective, and it does not generate any new data. The existing data present a risk of re-identification preventing its sharing according to approved IRB.

## ACKNOWLEDGEMENTS

This work was funded by grant no. UL1TR002550 from the National Center for Advancing Translational Sciences (NCATS) at the National Institutes of Health (NIH; E.J.T.), and by grant no. R21AG072349 from the National Institute on Aging (NIA) at the NIH (G.Q.).

## AUTHORS CONTRIBUTIONS

E.J.T., A.C.P., E.D.M., and G.Q., made substantial contributions to the study conception and design. A.C.P., M.G., and E.D.M. made substantial contributions to the preparation of data. F.M. implemented the model and conducted statistical analysis. F.M, A.C.P, E.D.M, and G.Q made substantial contributions to the interpretation of results. F.M and G.Q. drafted the first version of the manuscript. All authors contributed to critical revisions and approved the final version of the manuscript. E.D.M. and G.Q. take responsibility for the integrity of the work.

### COMPETING INTERESTS

The authors declare no competing interests.

## Notes

### Competing Interest Statement

The authors have declared no competing interest.

